# Convalescent plasma treatment of severe COVID-19: A matched control study

**DOI:** 10.1101/2020.05.20.20102236

**Authors:** Sean T. H. Liu, Hung-Mo Lin, Ian Baine, Ania Wajnberg, Jeffrey P. Gumprecht, Farah Rahman, Denise Rodriguez, Pranai Tandon, Adel Bassily-Marcus, Jeffrey Bander, Charles Sanky, Amy Dupper, Allen Zheng, Deena R. Altman, Benjamin K. Chen, Florian Krammer, Damodara Rao Mendu, Adolfo Firpo-Betancourt, Matthew A. Levin, Emilia Bagiella, Arturo Casadevall, Carlos Cordon-Cardo, Jeffrey S. Jhang, Suzanne A. Arinsburg, David L. Reich, Judith A. Aberg, Nicole M. Bouvier

**Author notes:** Drs. Aberg and Bouvier contributed equally to this article.

## Abstract

**Background:** Since December 2019, Coronavirus Disease 2019 (COVID-19) has become a global pandemic, causing mass morbidity and mortality. Prior studies in other respiratory infections suggest that convalescent plasma transfusion may offer benefit to some patients. Here, the outcomes of thirty-nine hospitalized patients with severe to life-threatening COVID-19 who received convalescent plasma transfusion were compared against a cohort of retrospectively matched controls.

**Methods:** Plasma recipients were selected based on supplemental oxygen needs at the time of enrollment and the time elapsed since the onset of symptoms. Recipients were transfused with convalescent plasma from donors with a SARS-CoV-2 (severe acute respiratory disease coronavirus 2) anti-spike antibody titer of ³1:320 dilution. Matched control patients were retrospectively identified within the electronic health record database. Supplemental oxygen requirements and survival were compared between plasma recipients and controls.

**Results:** Convalescent plasma recipients were more likely than control patients to remain the same or have improvements in their supplemental oxygen requirements by post-transfusion day 14, with an odds ratio of 0.86 (95% CI: 0.75∼0.98; p = 0.028). Plasma recipients also demonstrated improved survival, compared to control patients (log-rank test: p = 0.039). In a covariates-adjusted Cox model, convalescent plasma transfusion improved survival for non-intubated patients (hazard ratio 0.19 (95% CI: 0.05 ∼0.72); p = 0.015), but not for intubated patients (1.24 (0.33∼4.67); p = 0.752).

**Conclusions:** Convalescent plasma transfusion is a potentially efficacious treatment option for patients hospitalized with COVID-19; however, these data suggest that non-intubated patients may benefit more than those requiring mechanical ventilation.

## Introduction

Severe acute respiratory syndrome coronavirus 2 (SARS-CoV-2) is a positive-sense, single-stranded RNA virus belonging to the family *Coronaviridae*. Humans infected with SARS-CoV-2 may develop Coronavirus Disease 2019 (COVID-19), which manifests across a wide spectrum of clinical severity ranging from a mild upper respiratory tract illness to a diffuse viral pneumonia causing acute respiratory failure, with sequelae including acute lung injury, multi-organ dysfunction syndrome, and death.^1–3^ Antibody responses to coronavirus infections typically appear 2–3 weeks after the onset of illness and are rarely observed earlier.^4–6^

Although the relationship between disease severity and antibody response has yet to be firmly established,^7^ transfusion with convalescent plasma may provide a therapeutic option in the current treatment-limited environment.^8–10^ Historical evidence supports the efficacy of convalescent plasma transfusions to treat a variety of infectious diseases, including influenza, Junin virus, and severe acute respiratory syndrome (SARS).^11–14^ Initial data supporting convalescent plasma transfusions for COVID-19 include three case series from China of 5, 10, and 6 patients.^15–17^ In respiratory infections specifically, the strongest evidence suggests that the benefit of passive antibody transfer is most demonstrable in patients who were treated within days of symptom onset.^12,13,18,19^ Therefore, we hypothesized that treatment of patients with convalescent plasma early in the disease course may reduce morbidity and mortality associated with COVID-19. Presented here are preliminary outcomes for 39 patients with severe to life-threatening COVID-19 who received convalescent plasma transfusions at a single academic medical center, The Mount Sinai Hospital, in New York City.

## Methods

### Patients

Forty-five adult patients were identified as eligible for COVID-19 convalescent plasma transfusion under the criteria established for the FDA single patient emergency investigational new drug (eIND) process. FDA authorization was requested and obtained for COVID-19 convalescent plasma transfusion. Four patients improved and 2 patients withdrew consent prior to receipt of plasma, leaving 39 evaluable patients who received COVID-19 convalescent plasma. Patients were hospitalized in a single academic medical center in New York City for COVID-19 between 24 March 2020 and 8 April 2020. Patients were screened by symptom duration and by severity of disease on a case-by-case basis, as assessed by oxygen supplementation requirements and laboratory parameters. Patients or their legally authorized representatives provided informed consent prior to treatment. Both treatment and research were performed with the oversight of the Icahn School of Medicine at Mount Sinai Institutional Review Board (IRB).

### Convalescent plasma transfusion

Convalescent plasma donors were screened for SARS-CoV-2 antibody titers by a two-step Spike protein-directed ELISA.^20,21^ Donors with anti-spike antibody titers ≥1:320 were referred for blood collection at the New York Blood Center, which performed the plasmapheresis and then returned convalescent plasma units to The Mount Sinai Hospital. Plasma recipients were transfused with two units of ABO-type matched convalescent plasma. Each unit, approximately 250 milliliters in volume, was infused over 1 to 2 hours. Recipients were monitored every 15 minutes for signs of transfusion-related reactions and then followed post-transfusion for outcomes.

### Statistical analysis

To confirm the independent effect of convalescent plasma transfusion on improvement in oxygenation and survival, we conducted a propensity score-matched analysis using The Mount Sinai Hospital’s COVID-19 confirmed patient pool from the same calendar period (24 March 2020 to 8 April 2020). A logistic regression was fit to predict the potential for plasma therapy based on time series data obtained at baseline upon admission, prior to transfusion, and the day of transfusion. Among the predictors, exact matching was enforced on the administration of hydroxychloroquine and azithromycin, intubation status and duration, length of hospital stay, and oxygen requirement on the day of transfusion. Other medications were administered too infrequently to enforce exact matching. Balance was well achieved between the plasma and control groups, as all predictors had a standardized mean difference less than 0.2. Details of the matching method and results are described in the Supplementary Appendix. A medical data team reviewed charts of the control patients to determine outcomes at 1, 7, and 14 days. The data team was not informed of the recipient to whom each control patient was matched. Because control patients were matched to plasma recipients by length of stay prior to transfusion, “day 0” was defined as the day of transfusion for the plasma recipients and as the corresponding day in the hospitalization course of the control patients.

### Oxygen supplementation

Patients were then evaluated for their supplemental oxygen requirements and survival at three time points: days 1, 7, and 14 post-transfusion. Four categories of supplemental oxygen use status were collected for both cases and controls. These include, in order of increasing severity: room air without supplemental oxygen required; low-flow oxygen delivery by standard nasal cannula; high-flow oxygen delivery, including non-rebreather mask; high-flow nasal cannula or bi-level positive airway pressure (BiPAP) non-invasive ventilation; and mechanical ventilation. A patient’s oxygenation status at the three time points was considered to have worsened if they changed from a lower- to a higher-severity category compared to Day 0, or if they had died prior to the time point. A generalized estimating equations (GEE) approach with a logit link for binary data was used to model the effect of plasma on the odds of oxygenation improvement on days 1, 7, and 14 following transfusion, controlling for oxygen status on day 0. An independent working correlation structure was assumed for the patients within each cluster; however, the p-values were calculated based on the empirical standard errors. Since some patients were discharged with continued oxygen supplementation, the oxygen status of discharged patients was assumed to be no worse than low-flow oxygen by standard nasal cannula.

### Survival

Kaplan-Meier survival curves and the log rank test were used to depict the overall post-transfusion survival. A Cox model was fit to estimate the hazard ratio for in-hospital mortality for the plasma group, with matched clusters treated as random effects and onset of intubation as a time-varying covariate. In addition, interactions between convalescent plasma administration and intubation duration were tested to see if the plasma effects were the same in subgroups.

Both oxygen status and survival models were adjusted for duration of symptoms prior to admission and drugs administered, as these data were only ascertained after the matching was completed. The initial drug list consisted of COVID-19 therapies used during the time of the study that included azithromycin, broad-spectrum antibiotics, hydroxychloroquine, therapeutic anticoagulants, corticosteroids, directly acting antivirals, stem cells, and interleukin 1 and interleukin 6 inhibitors. Only those that had a p-value < 0.5, however, were included in the final model for adjustment. A liberal p-value was used here to be inclusive of any potential confounders. As a sensitivity analysis, the 1:2 matching without replacement data were also analyzed, where the balance between the matched pairs was enhanced but the study power was reduced. Descriptive data are reported as number (percent), mean (± standard deviation) or median [min, max], as appropriate. Analysis was performed using SAS 9.4 (SAS Institute Inc., Cary, NC). All tests were 2-sided and statistical significance was defined as a p value < 0.05, unless otherwise indicated.

## Results

### Recipient characteristics

The average age of the recipients of convalescent plasma transfusion was 55 (± 13) years (Table 1). The cohort was approximately two-thirds male and one-third female, similar to the proportions of men and women with severe disease in prior studies.^1^ Recipients generally had few baseline co-morbidities: 54% were obese (body mass index ≥30) and 18% had a current or former history of tobacco use. One patient had end-stage renal disease requiring peritoneal dialysis. The median duration of symptoms prior to initial presentation was 7 [0, 14] days. Inflammatory markers were elevated with median d-dimer of 1.4 [0.27, > 20] µg/mL fibrinogen equivalent units, median ferritin 1135 [107, 7441] ng/mL, and median C-reactive protein 159 [12, 319] mg/L. The median time between admission and transfusion was 4 [1, 7] days. On the day of transfusion, the majority of the recipients were requiring supplemental oxygen via a non-invasive delivery device (87%). Four plasma recipients (10%) were mechanically ventilated at the time of transfusion. In addition to receiving convalescent plasma transfusion, many recipients received a variety of inpatient pharmacotherapies throughout their hospitalizations (Table 2). There were no significant differences between plasma recipients and control patients in exposures to measured pharmacotherapies, except for therapeutic anticoagulation.

**Table 1.** Demographics and clinical parameters of recipients prior to transfusion.

| Characteristic* | Patients (N = 39) |
| --- | --- |
| **Demographics** |  |
| Mean age ± SD – year | 55 ± 13 |
| Sex – |  |
| Male / Female | 25 (64) / 14 (36) |
| Mean Body-mass index ± SD ‡ | 31.7 ± 6.0 |
| Coexisting disorder – no. (%) |  |
| Asthma | 3 (8) |
| Cancer¶ | 2 (5) |
| Chronic kidney disease | 1 (3) |
| Chronic obstructive pulmonary disease | 1 (3) |
| Current or former smoker | 7 (18) |
| Diabetes mellitus | 8 (21) |
| Hemorrhagic or ischemic stroke | 0 |
| Human immunodeficiency virus | 0 |
| Obesity | 21(54) |
| Obstructive sleep apnea | 2 (5) |
| Median duration of symptoms before admission – days | 7 [0, 14] |
| Presenting symptoms – no. (%) |  |
| Fever | 26 (67) |
| Shortness of breath | 26 (67) |
| Cough | 24 (62) |
| Diarrhea | 8 (21) |
| Sputum production | 3 (8) |
| Sore throat | 2 (5) |
| Vital signs on admission – no. (%) |  |
| Temperature >100.4°F or 38°C | 13 (33) |
| Heart rate >100 beats per min | 22 (56) |
| Respiratory rate ≥ 20 breaths per min | 28 (72) |
| Imaging – no. (%) |  |
| Chest radiography | 38 (97) |
| Chest computed tomography | 3 (8) |
| **Clinical parameters** |  |
| Laboratory data prior to transfusion |  |
| White-cell count |  |
| Median [range] – per mm<sup>3</sup> | 7600 [3900-22600] |
| Distribution – no (%) |  |
| ≥10,000/mm<sup>3</sup> | 10 (26) |
| ≤4000/mm<sup>3</sup> | 2 (5) |
| Aspartate aminotransferase >40 U/liter – no (%) | 26 (67) |
| Alanine aminotransferase >40/liter – no (%) | 18 (46) |
| Lactate ≥1.5 mmol/liter – no (%) | 23 (59) |
| D-dimer, median [range] - µg/ml Fibrinogen Equivalent units | 1.4 [0.27, >20] |
| Fibrinogen, mean (±S.D.) –no./total no. (mg/dl) | 684±140 |
| Ferritin, median [range] – ng/ml | 1135 [107, 7441] |
| C-Reactive Protein, median [range] – mg/liter | 159 [12, 319] |
| Interleukin-6, mean (±S.D.) – no./total no. (pg/ml) | 178±348 |
| Length of stay prior to transfusion |  |
| Median duration [range] – days | 4 [1, 7] |
| Supplemental oxygen requirement prior to initiation of transfusion |  |
| Standard nasal cannula – no. (%) | 7 (18) |
| 2 liters – no. (%) | 0 |
| 3 liters – no. (%) | 2 (5) |
| 4 liters – no. (%) | 2 (5) |
| ≥5 liters – no. (%) | 3 (8) |
| High-flow oxygen, high-flow nasal cannula or BiPAP – no. (%) | 27 (69) |
| Mechanical ventilation – no. (%) | 4 (10) |

\*Plus-minus values are mean ± SD. Percentages may not total 100 because of rounding.
‡The body-mass index is the weight in kilograms divided by the square of the height in meters.
¶Cancer represents a patient with thyroid cancer status post resection and a patient with Gleason 6 prostate cancer.
¶High-flow oxygen included venti-mask and non-rebreather mask; BiPAP = bi-level positive airway pressure.

**Table 2.** Recipient pharmacologic interventions.

|  |  | 1:4 matching | 1:2<br>matching |
| --- | --- | --- | --- |
|  | Patients | Controls | Controls |
| Pharmacologic interventions | (N = 39) | (N=156) | (N=74) |
| Antimicrobial agents – no. (%) |  |  |  |
| Azithromycin | 31 (79) | 133 (85) | 63 (85) |
| Broad spectrum antibiotics | 29 (74) | 112 (72) | 57 (77) |
| Hydroxychloroquine | 36 (92) | 148 (95) | 69 (93) |
| Investigational antivirals | 1 (3) | 9 (6) | 4 (5) |
| Therapeutic anticoagulation – no. (%) | 26 (67) | 64 (41) | 32 (43) |
| Anti-inflammatory agents – no. (%) |  |  |  |
| Corticosteroids | 22(56) | 90 (58) | 38 (51) |
| Interleukin-1 inhibitors | 0 | 0 | 0 |
| Interleukin-6 inhibitors | 3 (8) | 13 (8) | 6 (8) |

\* No significant differences were found between groups in both matched samples (p-values all >0.44), except for use of therapeutic anticoagulation (p <0.001 1:4 ratio and p=0.02 1:2 ratio).

### Respiratory Status

Plasma recipients and control patients were 100% matched on their supplemental oxygen requirement on day 0. Of them, 69.2% were receiving high-flow oxygen and 10.3% were receiving invasive mechanical ventilation. By day 14, clinical condition had worsened in 18.0% of the plasma patients and 24.3% in the control patients (p = 0.167, Cochran-Mantel-Haenszel test). The covariates-adjusted odds ratio for worsening oxygenation on day 14 was 0.86 (95% CI: 0.75∼0.98; p = 0.028) (Figure 1). The effect of plasma appeared to be confounded by the use of therapeutic anticoagulants (unadjusted vs. adjusted OR: 0.90 vs. 0.84), but not on other types of drugs or duration of symptoms before admission (OR remained in the range of 0.90∼0.91). On days 1 and 7, the plasma group also showed a reduction in the proportion of patients with worsened oxygenation status, but the group difference did not reach statistical significance.

**Figure 1.**
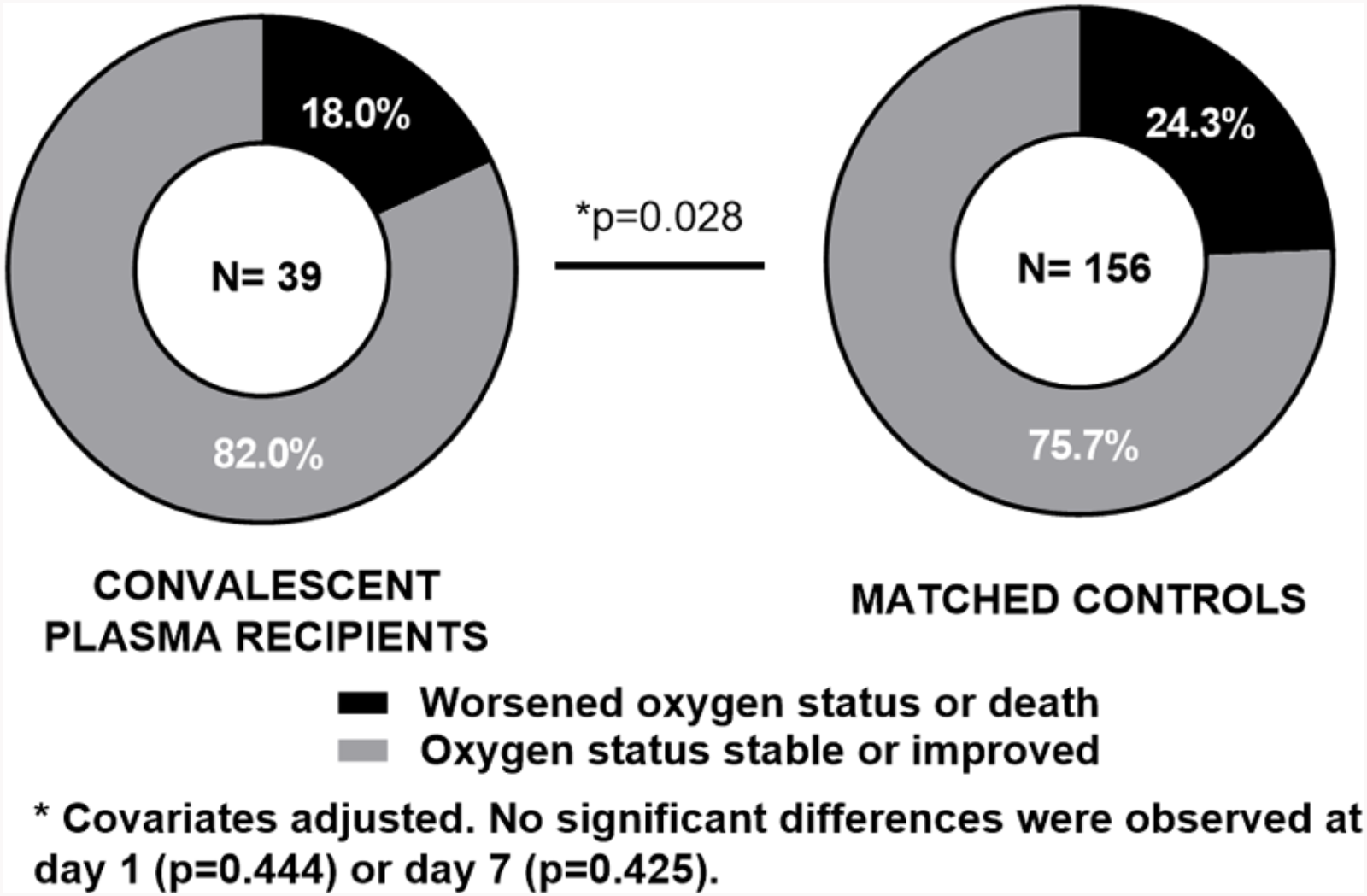
Comparison of oxygen requirements between Day 14 versus Day 0.

### Survival

As of 1 May 2020, 12.8% of plasma recipients and 24.4% of the 1:4 matched control patients had died (21.6% in the 1:2 matched dataset), and 71.8% and 66.7% (68.9%) had been discharged alive, respectively. The median follow-up time was 11 [1, 28] days for the plasma group and 9 [0, 31] days for the control group. Overall, we observed improved survival for the plasma group (log-rank test: p = 0.039) (Figure 2). In a covariates-adjusted Cox model, convalescent plasma transfusion was significantly associated with improved survival in non-intubated patients (hazard ratios: 0.19 (95% CI: 0.05 ∼0.72); p = 0.015), but not in intubated patients (1.24 (0.33∼4.67); p = 0.752) (P-value for the plasma and intubation interaction term was 0.050). There is no evidence that the effect of plasma depended on the duration of symptoms (p = 0.19 for the plasma by duration interaction). The results remain robust in the model without covariates adjustment and in the 1:2 matched sample (Figure 3).

**Figure 2.**
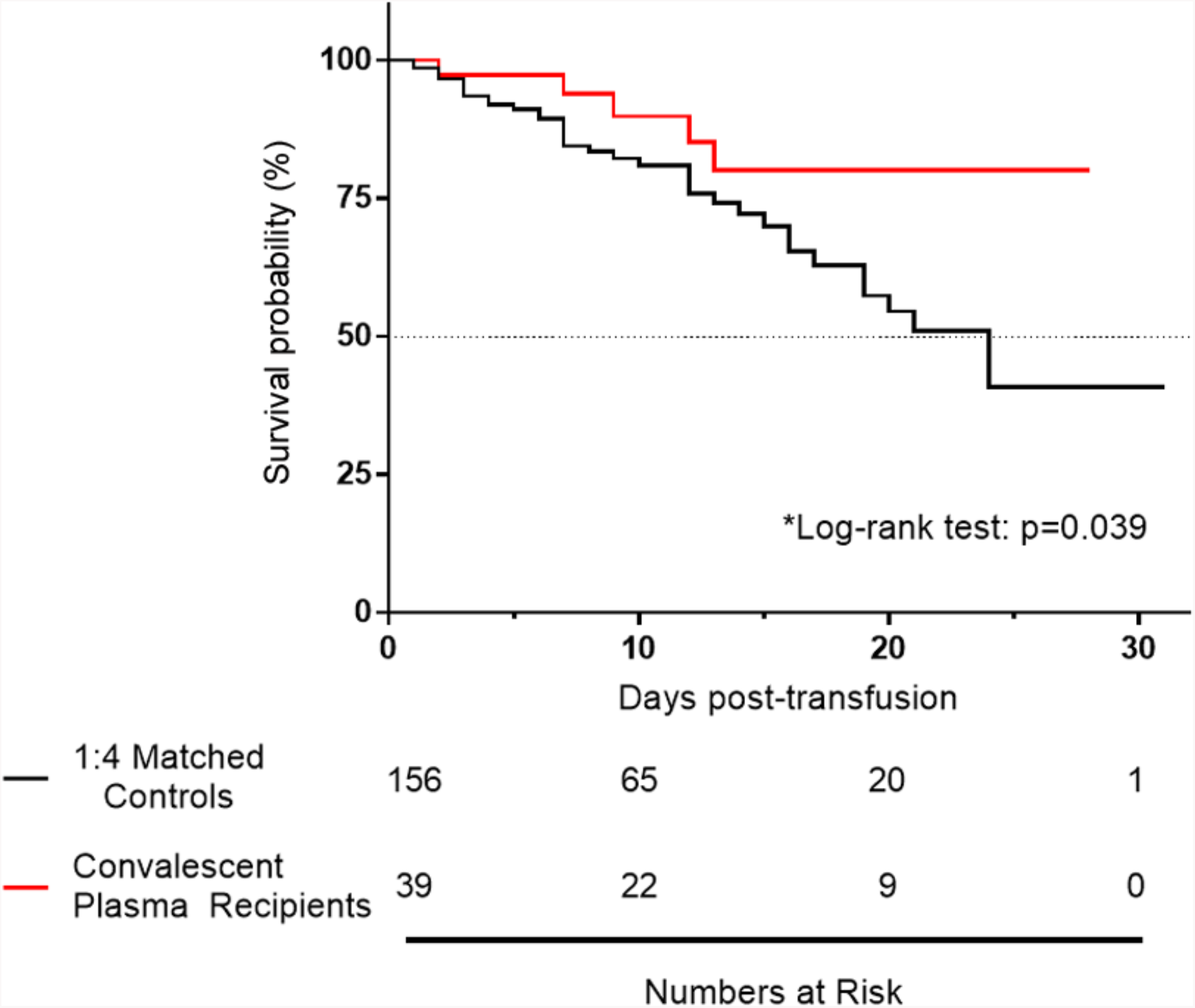
Survival Probability.

**Figure 3.**
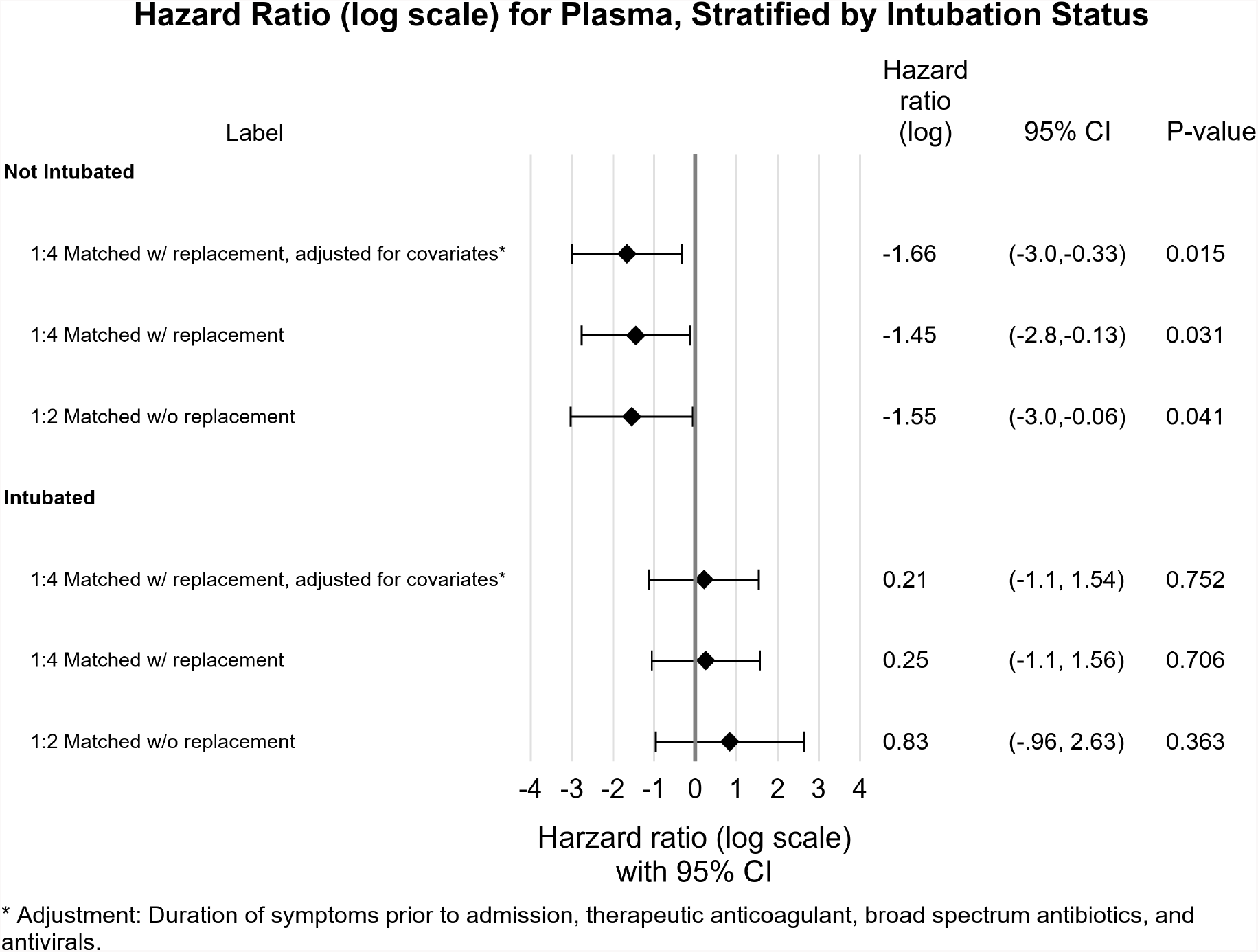
Hazard ratios for in-hospital mortality.

## Discussion

The COVID-19 pandemic poses an unprecedented challenge, as physicians and scientists struggle in real time to identify effective interventions against SARS-CoV-2 and its complications. This initial assessment offers evidence in support of convalescent plasma transfusion as an effective intervention in COVID-19. Preliminary data suggest a potential mortality benefit, but greater numbers are needed to draw definitive conclusions. Interestingly, these data suggest that the survival effect of convalescent plasma may begin to manifest more than 1 week after transfusion. If this observation is borne out in subsequent studies, it could indicate that convalescent plasma prevents longer-term complications, such as acute lung injury or multi-organ dysfunction syndrome; however, this speculation awaits confirmation in a larger patient cohort.

This study has many unique strengths. It describes the largest cohort of COVID-19 patients treated with convalescent plasma thus far worldwide. Furthermore, New York City has a large and very diverse population, and its metropolitan area was among the earliest and hardest hit by the COVID-19 pandemic in the United States. Over this study’s 16-day enrollment period (24 March 2020 to 8 April 2020), the Mount Sinai Health System admitted 4,152 confirmed COVID-19-positive patients. This large pool from which to draw control patients permitted an aggressive matching algorithm. Data from three different time frames –-baseline, prior to transfusion, and day of transfusion – informed the matching of controls to cases to maximize their similarity.

In addition, the efficacy of passive antibody transfer relies heavily on the quality of the donor convalescent plasma. Mount Sinai rapidly developed and clinically deployed an ELISA to titrate SARS-CoV-2-specific antibodies in serum,^18^ enabling our center to refer for blood collection only those convalescent donors with the highest peripheral serum antibody titers of ≥1:320.^22^ Prior smaller studies have reported on a variety of titer cutoffs,^15,16^ and at the time of this publication some centers are bypassing donor antibody titer pre-collection completely.^17^ Although the total quantity of anti-SARS-CoV-2 spike antibodies were assessed, it must be noted that we have not yet assessed the functionality of these antibodies in neutralizing the virus. Recent studies with SARS CoV-2 have generally found a high correlation between ELISA S protein binding activity and neutralization of SARS CoV-2.^21,23^

Although controls were retrospectively identified by propensity matching, the conclusions drawn from these data are not as robust as a prospective, randomized, placebo-controlled study. Furthermore, the convalescent plasma recipient cohort is highly heterogeneous in regards to oxygen needs at the time of transfusion and the duration of symptoms prior to admission. Other than intubated versus non-intubated patients, the small size of this cohort lacks sufficient power to permit additional subgroup analyses. We did not observe significant benefit of convalescent plasma in intubated patients, consistent with past literature demonstrating that passive antibody transfer therapies are most efficacious early in disease.^12,13,18,19^ However, the number of intubated patients in this study is small, limiting our ability to reach any conclusions about this population. Future studies that include more mechanically ventilated patients will be needed to address this uncertainty.

No significant transfusion-related morbidity or mortality were observed in the convalescent plasma recipient cohort; however, potential harms are associated with plasma transfusion. There is a risk of fluid volume overload, particularly in patients with end-stage renal disease or advanced heart failure. Allergic reactions to plasma are typically mild and self-limited. Plasma naturally contains procoagulants, whose additive effects are unknown in this disease, which is independently associated with hypercoagulability;^24^ thus, pending more data, additional caution should be exercised in patients with acute thrombotic events. Convalescent plasma transfusions also have theoretical risks, such as hindering the maturation of the patient’s own adaptive immune memory response and antibody-dependent enhancement. While keeping these risks in mind, additional studies are needed to confirm these findings and draw more definitive conclusions about the efficacy of convalescent plasma transfusion for the treatment of COVID-19 in different populations.

## Data Availability

Data are available from the corresponding author upon reasonable request.

## Acknowledgements

We thank all of the patients who participated in this study and their families. We also acknowledge the generosity of the thousands of anonymous tri-state area residents who recovered from COVID-19 and then volunteered to donate convalescent plasma for the benefit of others. We thank the New York Blood Center, Liise-anne Pirofski, Thomas Schneider, Carina Seah, Sindhu Srinivas, Douglas Tremblay, Freddy Nguyen, Miwa Geiger, Chaim Lebovits, and Jacqueline Lustgarten. We acknowledge the assistance of Icahn School of Medicine at Mount Sinai medical students: Sofia Ahsanuddin, Arence Paasewe, Ranjan Upadhyay, George Mellgard, Tyler Martinson, Bhavana Patil, Cynthia Luo, Saloni Agrawal, Alina Siddiqui, Julia Schwarz, Lydia Piendel, Jacqueline Emerson, Harrison Kaplan, Emma Klein, Mariely Garcia, James Johnson, Luke Maillie, and Elena Baldwin. We also appreciate the clinical expertise of the Mount Sinai Convalescent Plasma Squad: Nicholas Shuman, Daniela Delbeau, Donna Catamero, Gillian Sanchez, Suzan Aird, Manpreet Mann, Tarashon Broome, Sonia Kleiner-Arje, Louise Wolf, Angela Lee, Lisa Gaynes, and Karyn Goodman. We dedicate this work to the New Yorkers who have lost their lives to COVID-19 with a special dedication to the health care workers who will always be remembered for their selflessness during this pandemic.

## Author Affiliations

From the Division of Infectious Diseases (S.T.H.L., J.P.G., F.R., A.D., D.R.A., B.K.C., J.A.A., N.M.B.), and the Department of Population Health Science and Policy (H.-M.L., E.B.), and the Department of Pathology, Molecular and Cell-Based Medicine (I.B., D.R.M., A.F.B., C.C.-C., J.S.J., S.A.A.), and the Department of Medicine (A.W., J.B.), and The Tisch Cancer Institute (D.R.), and the Department of Surgical Critical Care (A.B.-M.), and the Department of Pulmonary and Critical Care Medicine (P.T.), and the Department of Anesthesiology, Perioperative and Pain Medicine (M.A.L., D.L.R.), and the Department of Medical Education (C.S.), and the Department of Microbiology (S.T.H.L., A.Z., F.K., N.M.B) – all at the Icahn School of Medicine at Mount Sinai; and the Department of Molecular Microbiology & Immunology, John Hopkins School of Medicine (A.C.).

## Disclosures

Dr. Krammer reports that patent applications have been filed for the assay used to select plasma donors, and Mount Sinai has licensed its use to several companies. Dr. Aberg reports grants and personal fees from Gilead, grants and personal fees from Merck, grants and personal fees from Janssen, personal fees from Theratech, personal fees from Medicure, grants from Regeneron, grants and personal fees from Viiv, outside the submitted work. All other authors have nothing to disclose. and the Department of Molecular Microbiology & Immunology, John Hopkins School of Medicine (A.C.).

## Notes

### Funding Statement

No external funding was received for this study.

